# *intI1* predicts ARGs and human source tracking markers carried by coprophagous flies in Maputo, Mozambique

**DOI:** 10.64898/2026.04.19.26351253

**Authors:** Anna A. Heintzman, Zaida Adriano Cumbe, Victoria Cumbane, Oliver Cumming, David Holcomb, Ishi Keenum, Jackie Knee, Vanessa Monteiro, Rassul Nalá, Joe Brown, Drew Capone

## Abstract

Wastewater surveillance is increasingly used for antimicrobial resistance (AMR) monitoring in urban environments, but low-resource settings often lack a piped sewerage system. Instead, coprophagous flies—flies that ingest feces—may serve as composite samplers for monitoring fecal wastes present in terrestrial environments. We evaluated whether the class 1 integron-integrase gene *intI1* was associated with genetic markers of AMR and fecal source tracking markers (FST) in coprophagous flies collected from latrine entrances and food preparation areas in low-income urban Maputo, Mozambique. We quantified *intI1*, an enteric 16S rRNA target (for normalization), three FST markers, and 30 ARG targets using qPCR. We normalized concentrations of *intI1* and each target to enteric 16S rRNA. We fit linear mixed models with a random intercept for housing compound to estimate within-fly associations between log_10_ relative abundance of *intI1* and log_10_ relative abundance of each target with and without adjustment for fly taxonomic group, capture location, and standardized fly mass. We also modeled per-fly unique ARG count (i.e., number of ARG targets detected) using Poisson regression. Of 188 flies assayed, 176 passed internal controls; *intI1* and enteric 16S rRNA were detected in 95% and 96% of flies, respectively. Higher relative abundance of *intI1* was positively associated with ARG and FST targets, with the strongest associations observed for sulfonamide-(*sul1*: β = 0.87; 95% CI: 0.81, 0.94; *sul2*: β = 0.81; 95% CI: 0.73, 0.89), tetracycline- (*tetA*: β = 0.78; 95% CI: 0.70, 0.85; *tetB*: β = 0.69; 95% CI: 0.60, 0.79), and trimethoprim-related (*dfrA17*: β = 0.78; 95% CI: 0.70, 0.86) genes. Associations with FST markers were weaker (i.e., human mtDNA: β = 0.46; 95% CI: 0.37, 0.55; human-associated *Bacteroides*: β = 0.34; 95% CI: 0.25, 0.43). Higher relative abundance of *intI1* was also associated with a greater number of ARGs detected: each 10-fold increase in *intI1* was associated with an 8% higher expected unique ARG count (aRR=1.08, 95% CI: 1.04–1.12). These findings support the need for further research across different settings exploring *intI1* carried by coprophagous flies as a potential standardized screening target for AMR surveillance in unsewered terrestrial environments.

## Introduction

Antimicrobial resistance (AMR) is a major and growing global public health threat driven by the emergence and spread of antimicrobial-resistant bacteria and the mobile genetic elements that facilitate the dissemination of resistance determinants (1,2). In 2019, bacterial AMR was estimated to be directly responsible for 1.27 million deaths and associated with 4.95 million deaths worldwide (3). Surveillance of environmental matrices is needed to further understand the localized and global burden of AMR (4,5). Low-resource settings typically have the least data available for decision making, but are where fecal wastes and other anthropogenic inputs are often ineffectively contained, resulting in frequent human exposure to contaminated domestic environments.

A persistent challenge for environmental AMR surveillance is the selection of molecular targets that are quantifiable, interpretable for health-relevant risk assessment, and informative about the potential for horizontal gene transfer. Standardization frameworks for AMR monitoring in environmental matrices have emphasized targets that reflect AMR-relevant contamination beyond overall bacterial biomass (6). Although sequencing-based approaches can provide detailed characterization of resistomes and mobile genetic elements, they may be impractical for routine monitoring in many settings because of cost, logistical requirements, and the sequencing depth often needed for low-biomass or dilute samples. Instead, standardizable qPCR targets may be more practical for these surveillance and screening applications. Within this context, the class 1 integron-integrase gene *intI1* is frequently included in qPCR panels because it is common in environments with anthropogenic pollution and is often linked to genetic elements conveying multidrug resistance (7,8). This “indicator target” approach is conceptually similar to the long-standing use of fecal indicator organisms (e.g., *E. coli*) in surface-water monitoring to infer broader fecal contamination and potential pathogen risk (9,10). Similarly, the World Health Organization Tricycle Program uses extended-spectrum beta-lactamase-producing (ESBL) *E. coli* as a standardized indicator for One Health AMR surveillance (4).

Class 1 integrons are genetic platforms that can capture and express gene cassettes, including many antibiotic resistance cassettes, and are frequently embedded within other mobile elements (e.g., transposons and plasmids) (7). The “clinical” class 1 integron lineage is thought to have been shaped by anthropogenic selective pressures and has been widely observed among bacteria in human- and domestic-animal-impacted environments. Consequently, *intI1* abundance has been proposed as a proxy for anthropogenic pollution, in part because of its linkage to antibiotic, disinfectant, and heavy-metal resistance determinants and its capacity to respond rapidly to environmental pressures through host growth and horizontal transfer (7).

Although much of the *intI1* literature has focused on aquatic environments, terrestrial near-household pathways may also be important for understanding AMR hazards in non-sewered settings with inadequate fecal waste containment. Synanthropic coprophagous flies are plausible mechanical vectors because they feed on feces and visit food and contact surfaces, thereby potentially transferring microbes and antibiotic resistance genes (ARGs) from waste sources into domestic environments. These behaviors make flies useful “composite samplers” that integrate microbial signals across multiple microenvironments that are difficult to capture through single-location sampling (11).

This study uses flies collected during the Maputo Sanitation (MapSan) trial, a controlled before-and-after evaluation of an onsite shared sanitation intervention in low-income informal neighborhoods of Maputo, Mozambique (12). Previous analyses of the MapSan trial demonstrated that flies collected in low-income neighborhoods of Maputo, Mozambique frequently carry enteric pathogen genetic targets and that an onsite shared sanitation intervention reduced fly counts at latrine entrances (13,14). In one analysis, the sanitation intervention was associated with reductions in ARG signals in flies, suggesting that sanitation infrastructure with basic fly-control features (e.g., pour flush toilets, netting on ventilation pipes) may limit fly-mediated dissemination of AMR-relevant genetic material.

In this study, we evaluate whether *intI1* measured in individual flies was associated with fly-associated AMR burden and fecal contamination. If *intI1* is consistently associated with multiple ARGs within individual flies, it may provide a standardizable, low-cost molecular approach to measure AMR in environmental matrices. Using gene copy measurements of *intI1*, ARGs, and fecal source tracking (FST) markers from individual flies collected during the Maputo Sanitation (MapSan) trial, we aimed to (i) estimate within-fly associations between *intI1* concentration and concentrations of prevalent ARG and FST targets and (ii) evaluate whether *intI1* is associated with the number of unique ARGs detected per fly as a summary measure of ARG diversity within the targeted panel.

## Materials and methods

### Maputo Sanitation trial

This analysis was conducted as a sub-study within the Maputo Sanitation (MapSan) trial, a controlled before-and-after (CBA) study designed to estimate health and exposure impacts of an urban onsite sanitation intervention in 16 low-income informal neighborhoods of Maputo, Mozambique (12). Study neighborhoods are characterized by poor housing and water, sanitation, and hygiene (WASH) conditions, a high burden of enteric disease, common indiscriminate antibiotic use, and high population density (approximately 15,000–25,000 persons/km²) (14–18). The intervention was delivered by a non-governmental organization (NGO) to household clusters (i.e., “compounds”) occupied by two or more households that shared sanitation and a common outdoor living space in which daily activities occurred (e.g., cooking, cleaning, and children’s play). Intervention sanitation systems (pour-flush toilets discharging to septic tanks with soakaway pits) were constructed within compound boundaries and therefore embedded in the immediate domestic living environment. The intervention infrastructure incorporated physical barriers intended to reduce vector contact with fecal wastes (*e.g.* mesh netting over ventilation pipes and water-seal toilets). Control compounds were enrolled concurrently from the same or adjacent neighborhoods and continued to use existing shared sanitation facilities throughout the study period. Greater detail regarding trial procedures, eligibility criteria, and intervention specifications is provided elsewhere (12,19).

### Data collection

Trained field enumerators visited enrolled intervention and control compounds at baseline (pre-intervention; April 1, 2015–March 31, 2016) and at the 12-month follow-up (April 1, 2016–March 31, 2017) (13,14). During household visits, enumerators obtained caregiver consent for participation in the broader MapSan trial and administered a caregiver survey on household demographics and behaviors (12). Enumerators then collected flies from latrine entrances and food preparation areas using paper traps coated with adhesive in 50 control and 50 intervention compounds at each study phase. Sticky traps were hung at least 1.5 m above ground and within 1 m of the latrine entrance and food preparation area (baseline: 5″ × 7″, Suterra, Bend, OR; 12-month: 5.5″ × 9.5″, Great Lakes IPM, Vestaburg, MI); the trap model was changed at follow-up due to manufacturer discontinuation. After 24 h of passive collection, field workers returned to enumerate the number of flies per trap and removed flies using tweezers sterilized with 10% bleach and 70% ethanol between compounds (but not between flies, to minimize time spent at households). All flies from a given trap were pooled into Whirl-Pak bags (Nasco, Fort Atkinson, WI) at baseline or sterile 15-mL centrifuge tubes (VWR, Radnor, PA) at 12 months, stored on ice, and transported to Ministry of Health laboratories in Maputo, Mozambique; samples were placed in a −80 °C freezer on the day of collection and shipped from Maputo to Atlanta, Georgia on dry ice with temperature monitoring for subsequent analysis. We used taxonomic keys to differentiate between house flies (*Musca domestica*) and blow flies (*Calliphoridae*), which are coprophagous, and other non-coprophagous flies (e.g. fruit flies, *Drosophilidae*), which we excluded from analysis (20).

### Nucleic acid extraction

Total nucleic acids were extracted from individual flies using a modified Qiagen DNeasy Blood & Tissue Kit (Qiagen, Hilden, Germany) with a pre-treatment step (21–23), following the procedures described in the prior MapSan fly analyses (13,14). We spiked each sample with two extraction positive controls and included at least one negative extraction control on each day of extractions (24).

### ARG and FST detection via custom TaqMan Array Card (TAC)

We assayed *intI1*, FST markers, and bacterial/ARG targets using a qPCR via a custom TaqMan Array Card (TAC) (ThermoFisher Scientific, Waltham, MA), as described in a previous MapSan manuscript (13). The TAC included an enteric 16S rRNA gene target, originally developed by Rousselon *et al.*, 2004 (25), designed to detect the Fec1 cluster of phylotypes, which has been reported to comprise approximately 5% of the human fecal microflora and was used here for normalization (25,26). The FST markers included on the card were bovine-associated *Bacteroides* DNA (BacCow), canine-associated *Bacteroides* DNA (BacCan), human-associated *Bacteroides* DNA (BacHum), and human mitochondrial DNA. The card also included a panel of 30 ARGs spanning eight antimicrobial classes that had previously been optimized for the TAC platform (S1 and S2 Tables) (27). Specifically, the ARG panel comprised genes conferring resistance to aminoglycosides (*armA*), chloramphenicols (*catA1*, *catB3*, *cmlA*, *floR*), colistin (*mcr-1*), fluoroquinolones (*gyrA83L, gyrA83S, parC80I, parC80S, qnrA, qnrB1*), two variants of aac(6′)-Ib that confer aminoglycoside and fluoroquinolone resistance (*aac6lb_104R, aac6lb_104W*), macrolides (*ermB*, *mphA*), tetracyclines (*tetA*, *tetB*), sulfonamides (*sul1, sul2*), trimethoprims (*dfrA17*), and β-lactams (*bla*_CTX-M1_*, bla*_CTX-M2/M74,_ *bla*_CTX-M8/M25_*, bla*_CTX-M9_*, bla*_NDM, OXA-1_*, bla*_OXA-9_*, bla_SHV_, bla*_VIM_). Quantitative PCR was performed on a QuantStudio 7 Flex instrument (ThermoFisher Scientific, Waltham, MA). For each run, we manually set the fluorescence threshold by comparing exponential amplification curves and multicomponent plots to positive control plots (S1 Fig), and we defined a target as detected only if amplification occurred before a quantification cycle (Cq) of 35.

### Ethics approval

Ethical approval for the MapSan Trial was obtained from the Mozambican Comité Nacional de Bioética para a Saúde (CNBS), Ministério da Saúde (333/CNBS/14) and the Georgia Institute of Technology Institutional Review Board (protocol #H15160). No additional permits were required for the work described in this study.

### Data analysis

#### Normalization and non-detect handling

Gene copy concentrations were log_10_-transformed prior to analysis. For primary models, we normalized all targets to bacterial load using the enteric 16S rRNA gene, expressing ARG and FST marker concentrations as log_10_ (target/16S rRNA) and *intI1* as log_10_ (*intI1*/16S rRNA).

Throughout the results, we refer to the 16S rRNA-normalized values as the relative abundance. Non-detects were imputed as one-half the theoretical limit of detection (83 gene copies per fly) before log transformation (S1 Text). Samples with non-detects for the 16S rRNA gene were excluded from 16S rRNA-normalized analyses.

#### Associations between *intI1* and individual targets (ARGs and FST markers)

To estimate within-fly associations between *intI1* and individual molecular targets, we *a priori* restricted regression analyses to ARG and FST markers with detection prevalence ≥20%. This threshold was selected to exclude low-prevalence targets for which the number of detections would limit statistical power to detect associations in linear regression models. We fit linear mixed models (LMMs) with log_10_ (ARG target/16S rRNA) as the outcome and log_10_ (*intI1*/16S rRNA) as the primary predictor. Models included a random intercept for compound to account for clustering of flies within compounds and were adjusted for covariates used in prior MapSan fly analyses, including fly taxonomic group (house fly vs. blow fly), capture location (latrine entrance vs. food preparation area), and fly mass (standardized to standard deviation from the mean) (13). We report effect estimates and 95% confidence intervals. We calculated Spearman rank correlations between *intI1* and each target and visualized key associations using scatter plots on the 16S rRNA-normalized scale, and repeated these analyses on the unnormalized scale to evaluate whether observed associations changed with variation in total bacterial load. Zhang *et al*. categorized ARGs into risk quartiles (Q1–Q4) based on a composite global health–risk score, with Q1 indicating the highest-risk genes (top 25% of ARGs with risk index > 0) and Q4 representing the lowest-risk (bottom 25% of ARGs with risk index > 0) (28). We report quartiles alongside Spearman rank correlations to contextualize whether *intI1* tracks ARGs prioritized as higher risk in a global ranking. Because our qPCR assays sometimes target gene families or mutation markers rather than single Antibiotic Resistance Ontology (ARO)-defined alleles, and because some targets are not included in Zhang *et al*.’s catalog, not all targets could be assigned a definite risk quartile.

#### Association between *intI1* and unique ARG count

We defined a per-fly “unique ARG count” as the number of ARG targets with positive detection in each sample to evaluate whether *intI1* was associated with the overall resistome. We modeled unique ARG count using a Poisson generalized linear mixed model (GLMM) with log_10_ (*intI1*/16S rRNA) as the primary predictor, a random intercept for compound, and adjustment for fly taxonomic group, capture location, and standardized fly mass. Exponentiated coefficients are reported as rate ratios (RR) and interpreted as the multiplicative change in expected unique ARG count per 10-fold increase in *intI1*/16S rRNA. Sensitivity analyses included alternative model specifications using unnormalized *intI1* gene copy concentrations, where rate ratios represent the multiplicative change in expected unique ARG count per 10-fold increase in *intI1* alone.

## Results

### Controls

Of the 188 flies assayed on the custom TaqMan Array Card, 12 were excluded because at least one extraction control failed to amplify as expected. All eight qPCR positive controls, which included plasmids with the primer/probe target sequences for each TAC assay (29), produced amplification for their respective targets. We observed no amplification across all targets in the 12 negative extraction controls.

### Fly prevalence and summary statistics

Among the 176 coprophagous flies included in the analysis, 159 (90%) were house flies (*Musca domestica)*, and 17 (10%) were green blowflies (*Calliphoridae).* Most flies were collected from food preparation areas (116/176; 66%), with the remainder caught at latrine entrances (60/176; 34%). Across all 176 flies, mean mass was 9.78 mg (SD 7.86), and median mass was 8.90 mg (IQR 4.80–12.67). For house flies, mean mass was 8.37 mg (SD 5.76), and median mass was 7.50 mg (IQR 4.20–11.80). For green blow flies, mean mass was 22.94 mg (SD 12.01), and median mass was 23.00 mg (IQR 12.50–28.20). Additional descriptive characteristics of the fly samples stratified by trial arm (control versus intervention), sampling phase (baseline versus 12-month follow-up), and collection location, as well as distributions of enteric 16S rRNA gene and ARG concentrations, have been reported previously (13).

### *IntI1*, ARG, and FST prevalence

Molecular detection of *intI1*, fecal source tracking markers, and ARG targets is summarized in Table 1. We detected the class 1 integron-integrase gene *intI1* in 96% (168/176) of flies and the enteric 16S rRNA gene in 96% (169/176). Among fecal source tracking markers, we detected a host-associated *Bacteroides* marker for human feces in 75% (132/176) of flies, followed by a host-associated *Bacteroides* marker for canine feces in 35% (62/176), and a host-associated *Bacteroides* marker for bovine feces in 18% (32/176). We detected human mtDNA, a human fecal source tracking marker, in 59% (103/176) of flies.

**Table 1.**
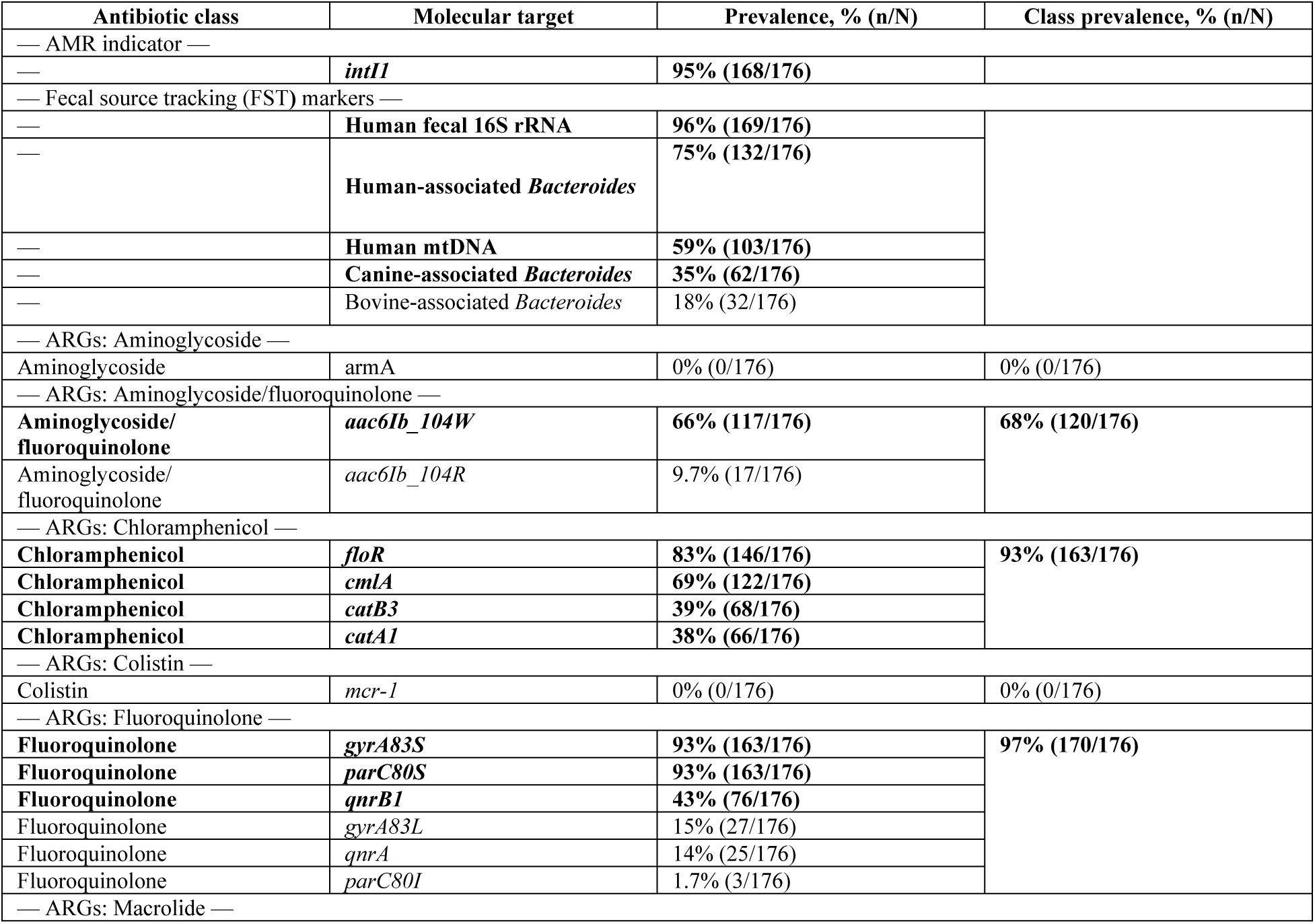
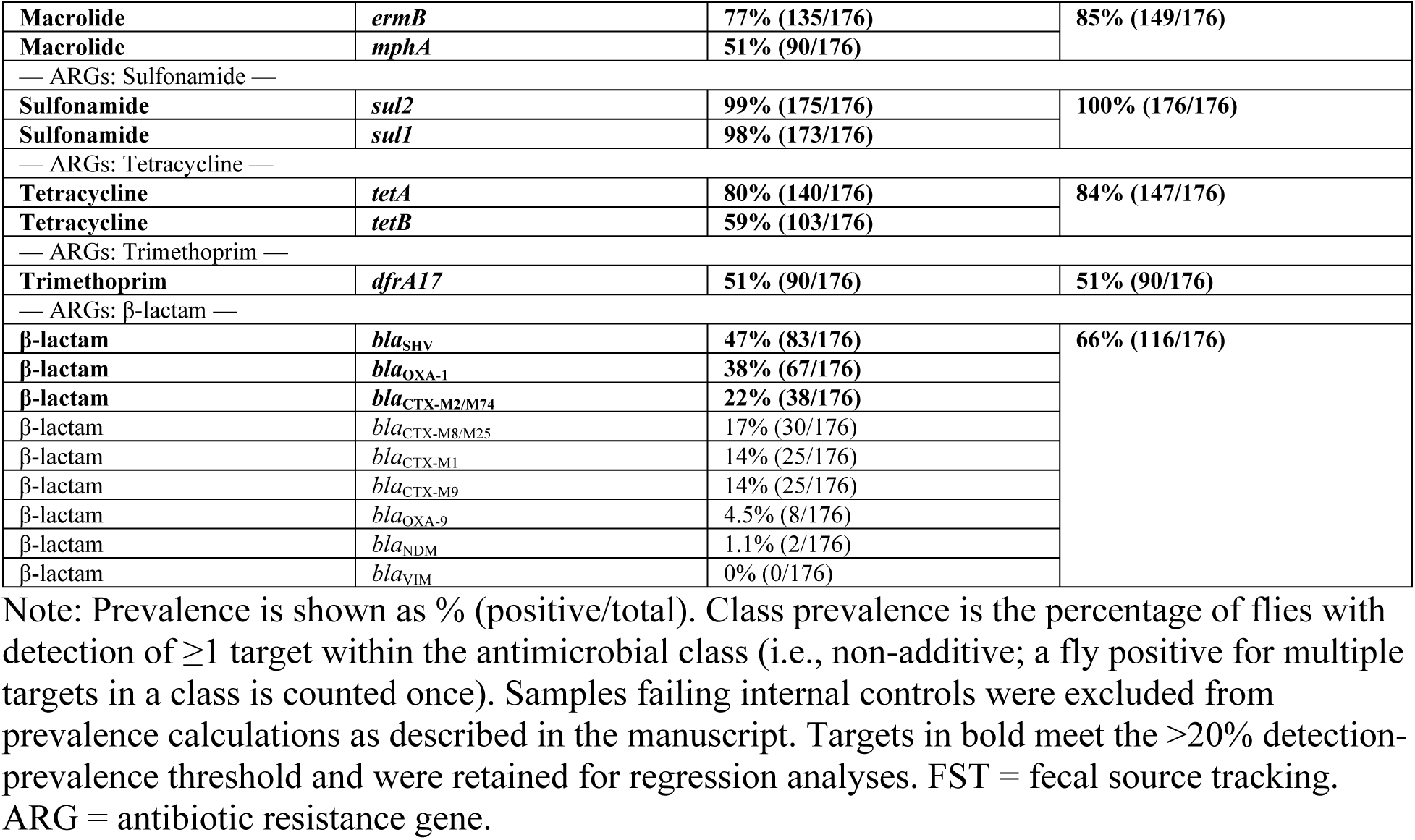
Detection prevalence of *intI1*, fecal source tracking markers, and ARG targets across fly samples (n = 176) collected in Maputo, Mozambique.

We detected at least one sulfonamide resistance gene in 100% (176/176) of flies, including *sul2* in 99% (175/176) and *sul1* in 98% (173/176). We detected at least one fluoroquinolone-associated target in 97% (170/176) of flies; within this class, *gyrA83S* and *parC80S* were each detected in 93% (163/176), followed by *qnrB1* in 43% (76/176). We detected other fluoroquinolone-associated targets less frequently, including *gyrA83L* in 15% (27/176), *qnrA* in 14% (25/176), and *parC80I* in 1.7% (3/176). We detected chloramphenicol resistance genes in 93% (163/176) of flies, with *floR* detected in 83% (146/176) and *cmlA* in 69% (122/176); *catB3* and *catA1* were detected in 39% (68/176) and 38% (66/176), respectively. We detected macrolide resistance genes in 85% (149/176) of flies, including *ermB* in 77% (135/176) and *mphA* in 51% (90/176). We detected tetracycline resistance genes in 84% (147/176) of flies, including *tetA* in 80% (140/176) and *tetB* in 59% (103/176). We detected at least one of the two genes conferring aminoglycoside/fluoroquinolone resistance in 68% (120/176) of flies; 66% (117/176) and 9.7% (17/176) of flies were positive for *aac6Ib_104W* and *aac6Ib_104R*, respectively. We detected at least one β-lactam resistance gene in 66% (116/176) of flies; *bla*_SHV_ and *bla*_OXA-1_ were most prevalent in 47% (83/176) and 38% (67/176), respectively, while *bla*_CTX-M_ targets were less common: *bla*_CTX-M2/M74_ in 22% (38/176), *bla*_CTX-M8/M25_ in 17% (30/176), *bla*_CTX-M1_ in 14% (25/176), and *bla*_CTX-M9_ 14% in (25/176). We detected *bla*_OXA-9_ in 4.5% (8/176) and *bla*_NDM_ in 1.1% (2/176), and we did not detect *bla*_VIM_ (0%, 0/176). We detected the trimethoprim resistance gene *dfrA17* in 51% (90/176). We did not detect *armA* (0%, 0/176) or *mcr-1* (0%, 0/176).

### Associations between intI1 and individual targets

We observed consistently positive associations between *intI1* and both FST markers and ARG targets. In the primary (16S rRNA-normalized) models, we interpreted β as the expected change in the log_10_ relative target abundance per 1 log_10_-increase in relative *intI1* abundance (Table 2, Fig 1). Across most targets, adjustment for fly taxonomic group, capture location, and standardized fly mass produced effect estimates similar to unadjusted estimates.

**Fig 1.**
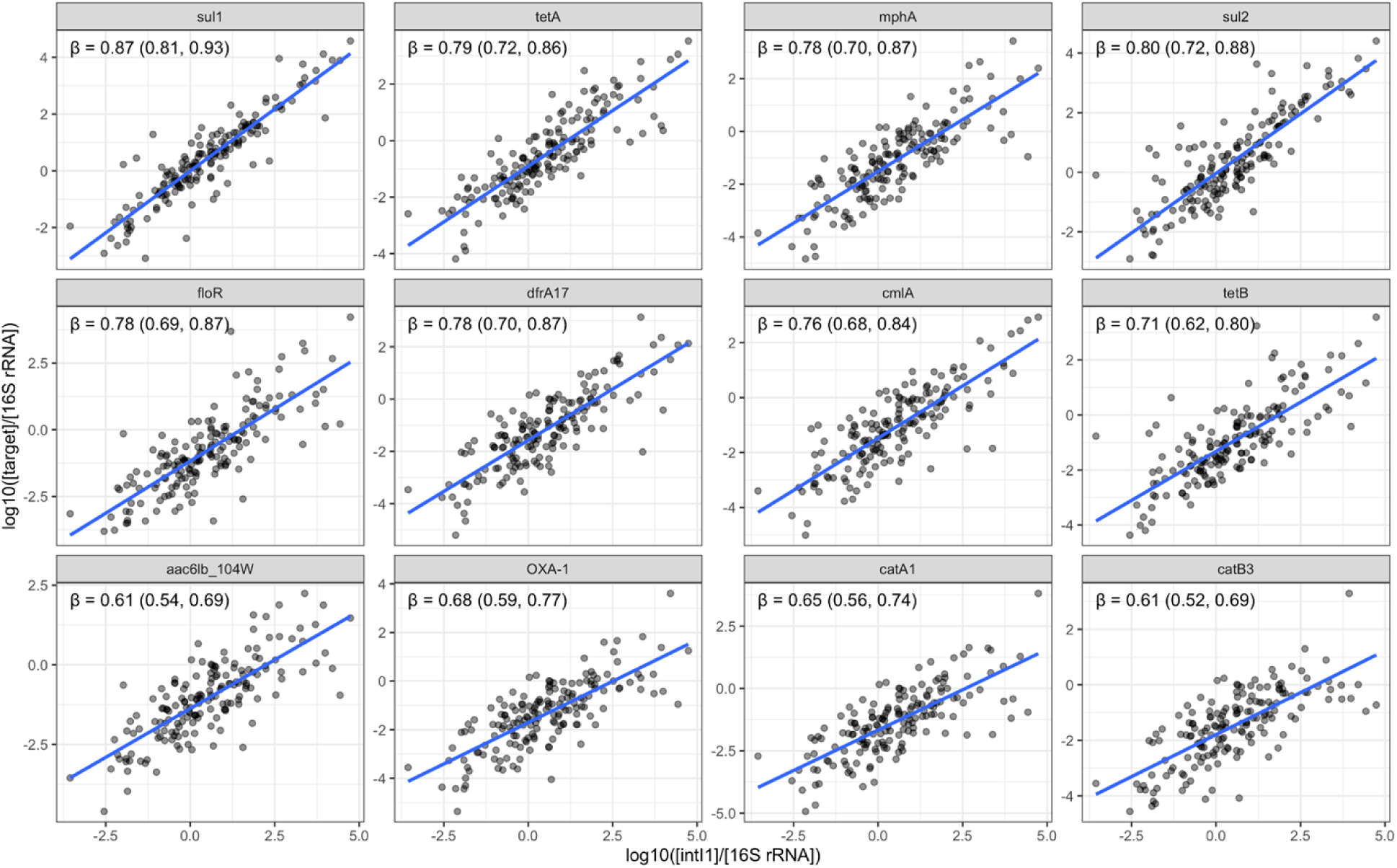
Associations between *intI1* (16S rRNA-normalized) and the most strongly correlated ARG and FST targets (16S rRNA-normalized) in flies collected in Maputo, Mozambique. Faceted scatterplots show log_10_(target/16S rRNA) versus log_10_(*intI1*/16S rRNA) for the 12 targets with the strongest correlations. Points represent individual fly samples. Blue lines show fixed-effect predictions from a linear mixed-effects model with a random intercept for compound. The annotated β (95% CI) is the fixed-effect slope for the association between log_10_(*intI1*/16S rRNA) and log_10_(target/16S rRNA) within each facet. FST = fecal source tracking. ARG = antibiotic resistance gene.

**Table 2.**
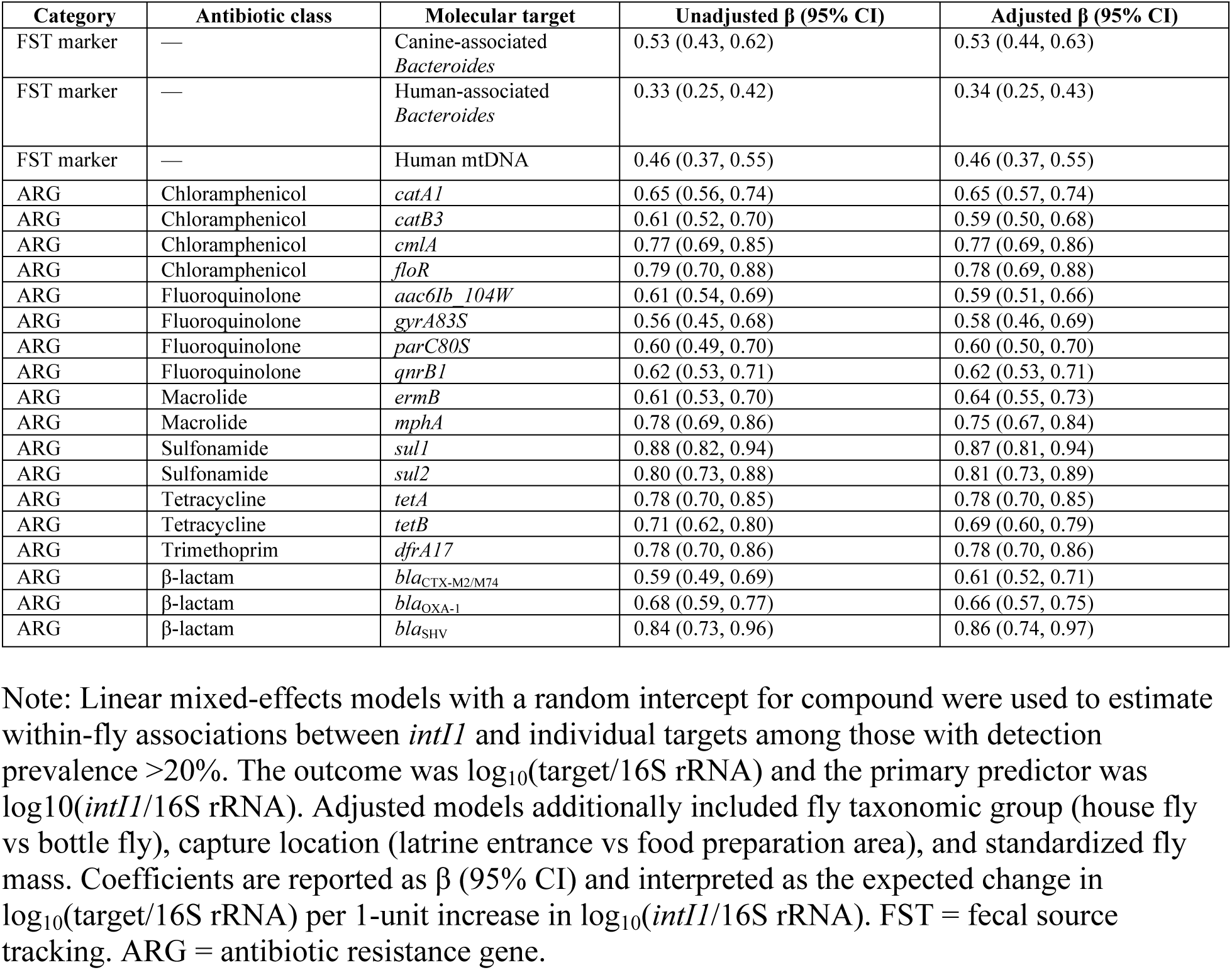
Associations between *intI1* (16S rRNA-normalized) and FST markers and ARG targets (16S rRNA-normalized) in flies collected in Maputo, Mozambique.

We observed positive 16S rRNA-normalized associations between *intI1* and all three retained FST markers, including the host-associated *Bacteroides* marker for canine fecal pollution (β = 0.53; 95% CI: 0.44, 0.63), the host-associated *Bacteroides* marker for human fecal pollution (β = 0.34; 95% CI: 0.25, 0.43), and human mtDNA (β = 0.46; 95% CI: 0.37, 0.55) in adjusted models.

Across ARG targets, the log_10_ relative *intI1* abundance was positively associated with targets spanning multiple antimicrobial classes. For sulfonamide resistance genes, a 1 log_10_-increase in the relative *intI1* abundance was associated with 0.87 log_10_ higher relative *sul1* abundance (β = 0.87; 95% CI: 0.81, 0.94) and 0.81 log_10_ higher relative *sul2* abundance (β = 0.81; 95% CI: 0.73, 0.89) in adjusted models. For tetracycline targets, a 1 log_10_-increase in the relative *intI1* abundance was associated with 0.78 log_10_ higher relative *tetA* abundance (β = 0.78; 95% CI: 0.70, 0.85) and 0.69 log_10_ higher relative *tetB* abundance (β = 0.69; 95% CI: 0.60, 0.79) (Table 2). For macrolide targets, a 1 log_10_-increase in the relative *intI1* abundance was associated with 0.64 log_10_ higher relative *ermB* abundance (β = 0.64; 95% CI: 0.55, 0.73) and 0.75 log_10_ higher relative *mphA* abundance (β = 0.75; 95% CI: 0.67, 0.84).

Within fluoroquinolone-associated targets, a 1 log_10_-increase in the relative *intI1* abundance was associated with 0.59 log_10_ higher relative *aac6Ib_104W* abundance (β = 0.59; 95% CI: 0.51, 0.66), 0.58 log_10_ higher relative *gyrA83S* abundance (β = 0.58; 95% CI: 0.46, 0.69), 0.60 log_10_ higher relative *parC80S* abundance (β = 0.60; 95% CI: 0.50, 0.70), and 0.62 log_10_ higher relative *qnrB1* abundance (β = 0.62; 95% CI: 0.53, 0.71) in adjusted models (Table 2). For chloramphenicol targets, a 1 log_10_-increase in the relative *intI1* abundance was associated with 0.65 log_10_ higher relative *catA1* abundance (β = 0.65; 95% CI: 0.57, 0.74), 0.59 log_10_ higher relative *catB3* abundance (β = 0.59; 95% CI: 0.50, 0.68), 0.77 log_10_ higher relative *cmlA* abundance (β = 0.77; 95% CI: 0.69, 0.86), and 0.78 log_10_ higher relative *floR* abundance (β = 0.78; 95% CI: 0.69, 0.88) (Table 2). For β-lactam targets, a 1 log_10_-increase in the relative *intI1* abundance was associated with 0.61 log_10_ higher relative *bla*_CTX-M2/M74_ abundance (β = 0.61; 95% CI: 0.52, 0.71), 0.66 log_10_ higher relative *bla*_OXA-1_ abundance (β = 0.66; 95% CI: 0.57, 0.75), and 0.86 log_10_ higher relative *bla*_SHV_ abundance (β = 0.86; 95% CI: 0.74, 0.97).

In unnormalized models, we interpreted β as the expected change in log_10_(target) per 1 log_10_-increase in *intI1* gene copies, corresponding to a 10-fold increase in *intI1* (S3 Table, S2 Fig). In these models, higher log_10_(*intI1*) was associated with higher log_10_(16S rRNA) (adjusted β = 0.24; 95% CI: 0.09, 0.39), indicating that *intI1* co-occurred with the bacterial indicator for overall fecal load. Unnormalized associations between *intI1* and individual targets remained positive across retained FST markers and ARG targets, with effect estimates generally attenuated relative to 16S rRNA-normalized models but similar in direction and robust to covariate adjustment.

To compare the relative strength of *intI1*–target relationships across retained targets (>20% prevalence), we ranked Spearman correlations between the log_10_ relative *intI1* abundance and log_10_ relative target abundance (Fig 2). Among ARGs with Zhang *et al*. risk-quartile assignments (28), the strongest correlations overall were observed for several Q1 targets, including *sul1* (ρ = 0.90), *tetA* (ρ = 0.87), *mphA* (ρ = 0.83), *sul2* (ρ = 0.81), *ermB* (ρ = 0.71), and *OXA-1* (ρ = 0.75).

**Fig 2.**
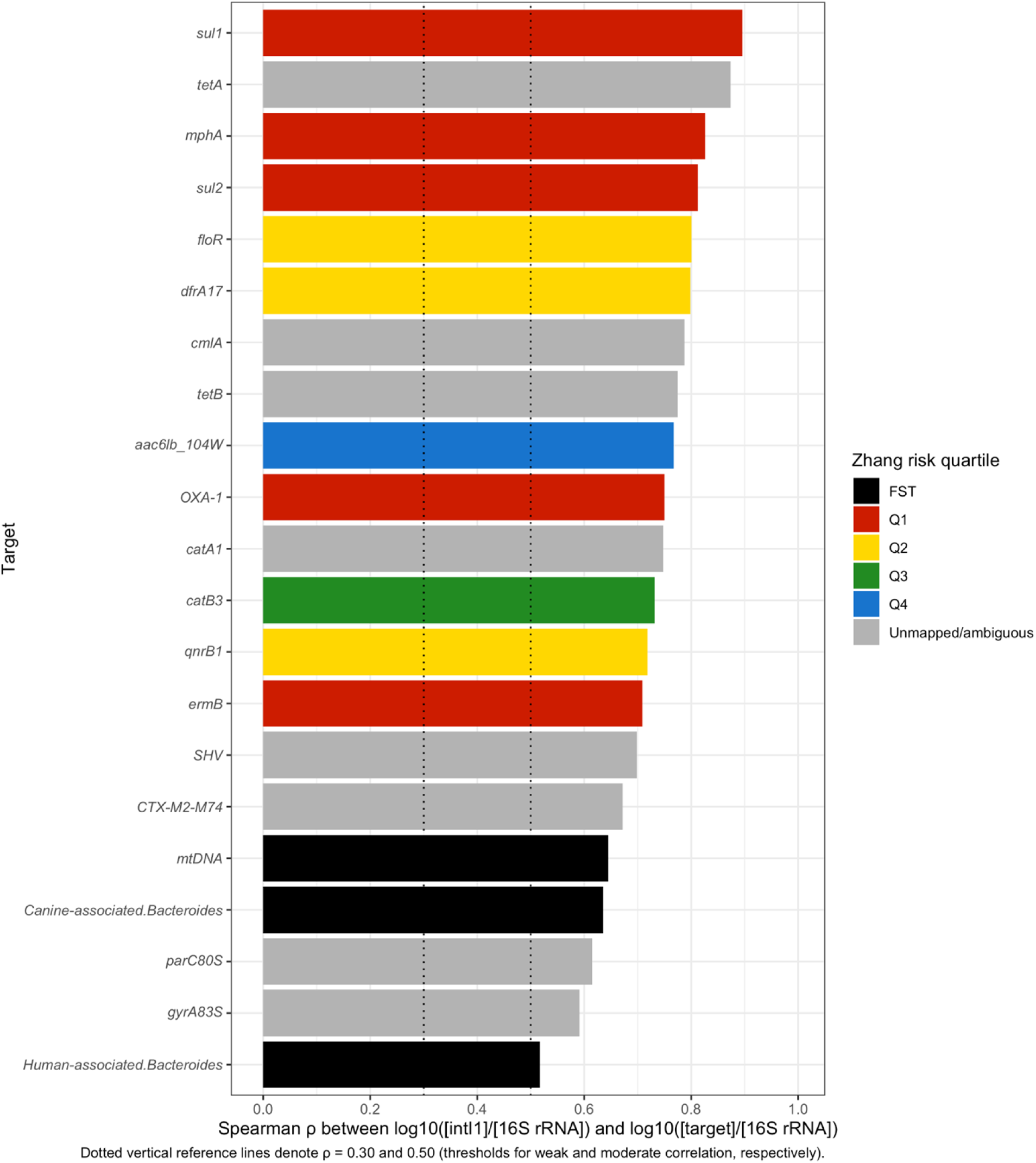
Ranked Spearman correlations between *intI1* (16S rRNA-normalized) and ARG/FST targets (16S rRNA-normalized) in flies collected in Maputo, Mozambique. Horizontal bars show Spearman rank correlation coefficients (ρ) between log_10_(*intI1*/16S rRNA) and log_10_(target/16S rRNA) across fly samples. Targets are ordered from highest to lowest ρ. Bar colors indicate Zhang *et al*. ARG risk quartile assignment (Q1–Q4), where Q1 indicates the highest-risk genes (top 25% of ARGs with risk index > 0) and Q4 represents the lowest-risk (bottom 25% of ARGs with risk index > 0) (28). FST markers are shown in black; targets without a risk-quartile mapping are shown in gray (unmapped/ambiguous). Dotted vertical reference lines denote ρ = 0.30 and 0.50 (thresholds for weak and moderate correlation, respectively). FST = Fecal Source Tracking. ARG = antibiotic resistance gene.

Within Q2, correlations remained high for a few targets (*floR*, ρ = 0.80; *dfrA17*, ρ = 0.80; *qnrB1*, ρ = 0.72). Q3 targets were also strongly correlated, including *catB3* (ρ = 0.73) and the β-lactam group *bla*_CTX-M2/M74_ (ρ = 0.67). The single Q4 target (*aac6Ib_104W*) showed a strong correlation with *intI1* (ρ = 0.77). Several targets with ambiguous Zhang mappings also correlated positively with *intI1*, including *SHV* (ρ = 0.70), *cmlA* (ρ = 0.79), *catA1* (ρ = 0.75), and the fluoroquinolone-associated mutation markers *parC80S* (ρ = 0.62) and *gyrA83S* (ρ = 0.59). In contrast, FST markers showed more moderate correlations with *intI1*, including human mtDNA (ρ = 0.65), the canine-associated *Bacteroides* marker (ρ = 0.64), and the human-associated *Bacteroides* marker (ρ = 0.52). Unnormalized correlations showed the same overall direction but were generally attenuated (Fig S3).

### Association between *intI1* and unique ARG count

Across fly samples, higher relative *intI1* abundance was associated with a greater number of unique ARG targets detected (Figs 3 and 4). In the unadjusted model, a 10-fold increase in the relative *intI1* abundance was associated with a 6% higher expected unique ARG count (RR = 1.06, 95% CI: 1.03–1.10); the association was similar after adjustment for fly taxonomic group, capture location, and standardized fly mass (RR = 1.08, 95% CI: 1.04–1.12) (Fig 3). Observed counts varied substantially at a given log_10_ relative *intI1* abundance value, but model-predicted mean counts increased monotonically in both models, with slightly steeper increases in the adjusted model (Fig 4). In the unnormalized models, higher *intI1* was more strongly associated with a greater number of unique ARG targets (S4 Fig and S5 Fig).

**Fig 3.**
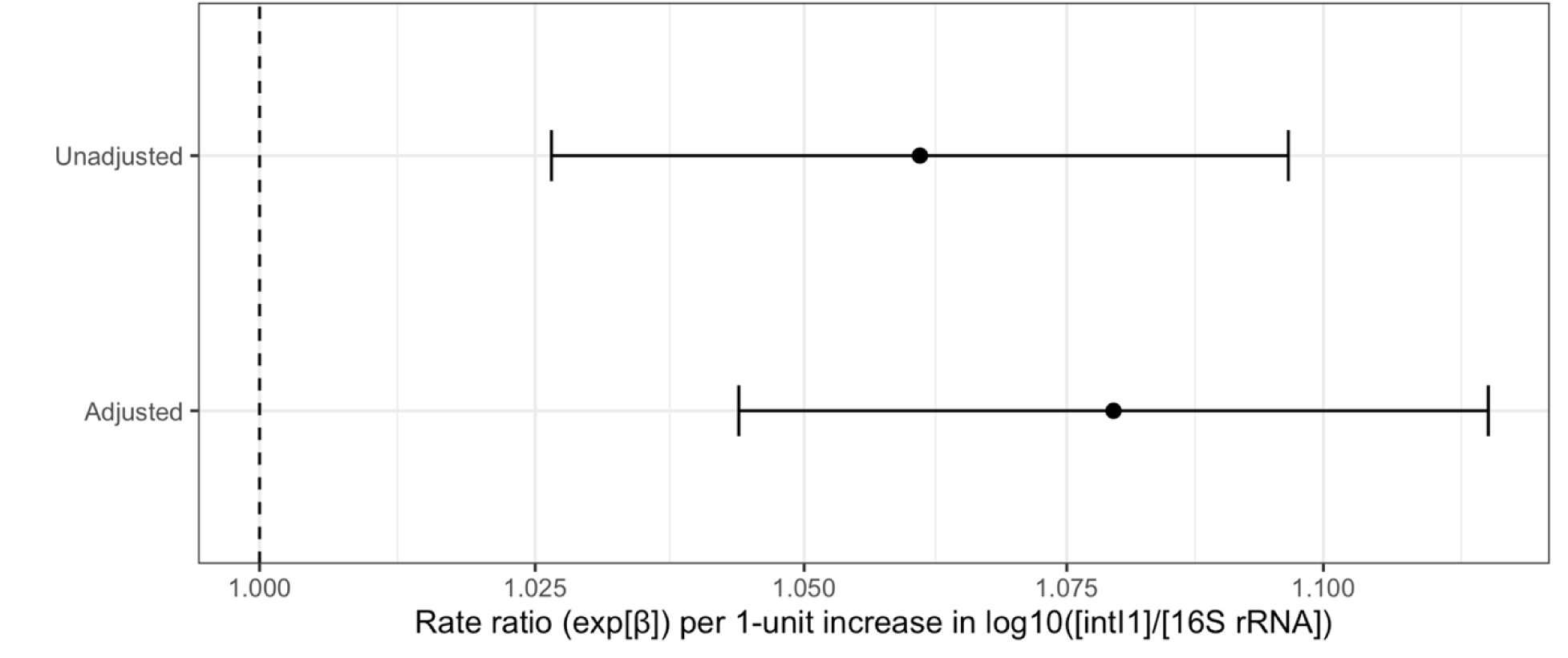
Association between *intI1* (16S rRNA-normalized) and unique ARG count in flies collected in Maputo, Mozambique. Forest plot of exponentiated coefficients (rate ratios; RR) from Poisson generalized linear mixed models (GLMMs) relating unique ARG count (number of ARG targets detected per fly) to log_10_(*intI1*/16S rRNA). Points denote RR estimates and horizontal lines show 95% confidence intervals for unadjusted and adjusted models. Both models include a random intercept for compound; the adjusted model additionally includes fly taxonomic group, capture location, and standardized fly mass. Rate ratios are interpreted as the multiplicative change in expected unique ARG count per 1-unit increase in log_10_(*intI1*/16S rRNA). The dashed vertical line indicates RR = 1.0 (no association). Samples with non-detect 16S rRNA were excluded. ARG = antibiotic resistance gene.

**Fig 4.**
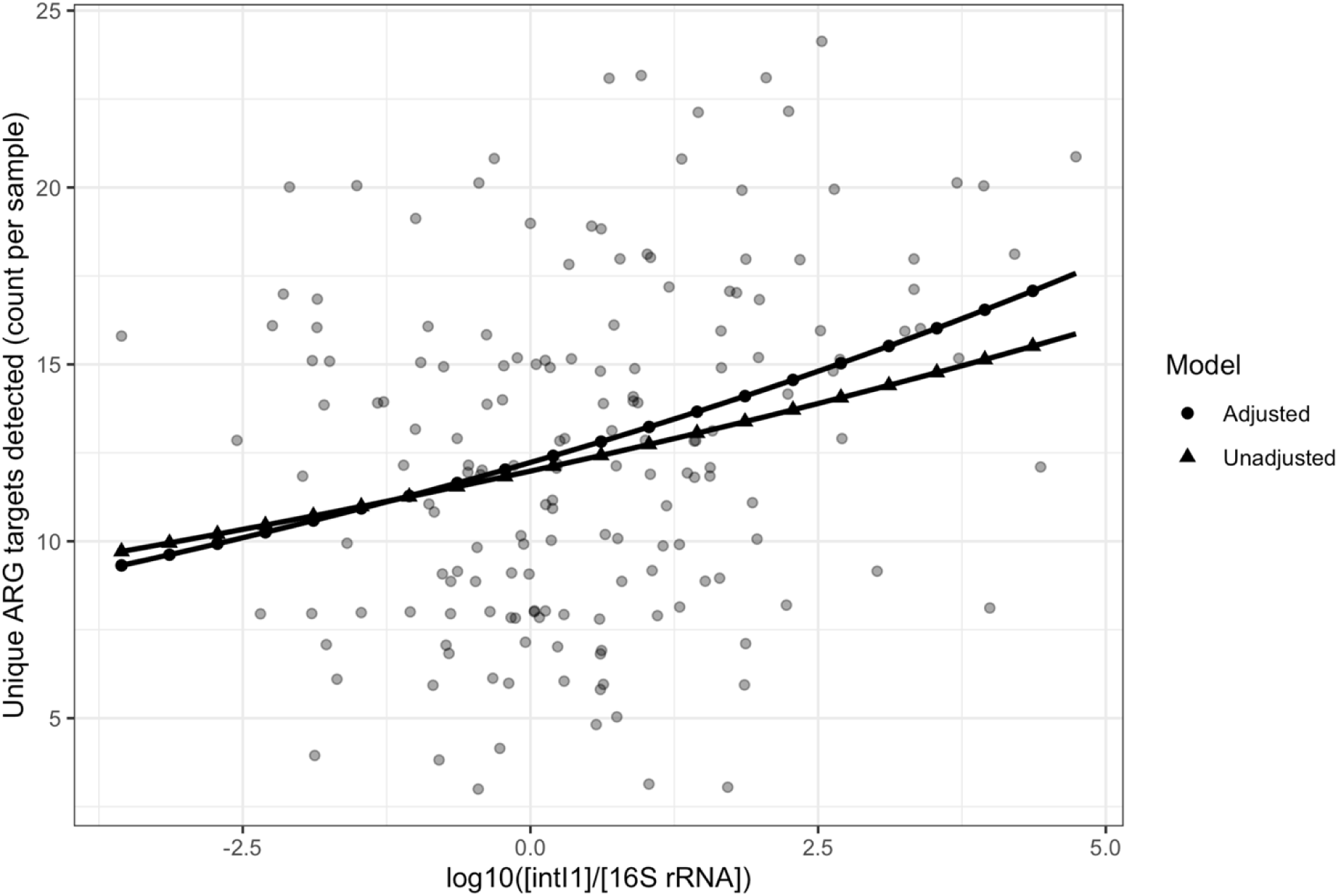
Observed and model-predicted unique ARG count given *intI1* (16S rRNA-normalized) in flies collected in Maputo, Mozambique. Scatterplot of observed unique ARG count per fly versus log_10_(*intI1*/16S rRNA) with overlaid model-predicted mean counts from Poisson GLMMs. Points represent individual fly samples. Lines show predicted mean unique ARG count from the unadjusted model (random intercept for compound only) and the adjusted model (random intercept for compound plus fly taxonomic group, capture location, and standardized fly mass). Predictions are shown across the observed range of log_10_(*intI1*/16S rRNA). ARG = antibiotic resistance gene.

## Discussion

In flies collected in Maputo, Mozambique, *intI1* was an indicator of individual ARGs and an indicator of the unique number of ARGs in each sample. Higher 16S rRNA-normalized *intI1* was associated with higher 16S rRNA-normalized concentrations of all retained ARG targets spanning multiple antimicrobial classes, with similar estimates in adjusted and unadjusted models. Consistent with these results, Spearman correlations between the relative *intI1* abundance and the relative target abundance were strongest for several sulfonamide, tetracycline, and trimethoprim targets, and correlations remained positive in all other retained ARGs. The relative *intI1* abundance was also associated with a higher unique ARG count per fly, supporting its potential use as a single-marker proxy for AMR carried by coprophagous flies. These findings align with prior environmental studies that treat *intI1* as an indicator of anthropogenic AMR pollution (7), and extend that concept to coprophagous flies, which can act as “composite samplers” of fecal material in unsewered settings.

Although *intI1* was positively associated with fecal source tracking (FST) markers, correlations with FST markers were generally weaker than correlations with many ARG targets. This pattern suggests that the fly-associated AMR signal captured by *intI1* is not simply a function of fecal-source loading and may not be consistently linked to a single animal or human source, which is potentially different than wastewater sampling from sewered systems. This may also reflect the local animal profile and mixed exposure environment in Maputo. In baseline MapSan household surveys, 7.7% of households reported a dog present, and latrines were typically shared across 3–5 households (18). Low dog ownership could plausibly limit canine fecal inputs and help explain the comparatively modest association with the canine-associated *Bacteroides* marker.

Additionally, there were no cows in the study area, so positive detection of the bovine-associated marker should not necessarily be interpreted as evidence of bovine fecal contamination. BacCow, the bovine-associated *Bacteroides* target we used in our assay, has been described as a broader livestock- or non-human-associated *Bacteroidales* marker rather than a cow-exclusive target(30). In the parent trial, compounds had a mean population of 14–19 residents, depending on arm/phase. The greater population density of humans than animals, likely contributed to the larger association between *intI1* and human FST markers (31).

These findings suggest that *intI1* may be a useful indicator of ARG burden in individual flies in low-resource, unsewered settings where flies frequently contact fecal material. More broadly, the results support evaluating coprophagous flies as potential composite samples for AMR surveillance in environments with inadequate fecal waste containment.

This study has several limitations. First, qPCR non-detect handling required substitution for *intI1* non-detects and exclusion of 16S rRNA non-detects in normalized analyses, which may influence estimates near the detection limit. Second, the observational association framework cannot establish directionality or physical linkage between *intI1* and specific ARGs; further analysis via sequencing is necessary to determine if *intI1* is on the same gene cassette as these ARGs. Third, sampling constraints from the fly collections (including changes in trap models across phases) may introduce variability not fully captured in the models.

Overall, *intI1* measured in individual flies was strongly and consistently associated with multiple ARG targets and with the number of unique ARGs detected per fly, supporting its utility as a sentinel marker for fly-associated AMR burden. If validated in other environmental contexts, *intI1* could serve as a low-cost screening target for fly-based surveillance, potentially paired with a focused panel of higher-priority ARGs. Future work should evaluate whether *intI1* is also a correlate of enteric pathogen targets in flies and use sequencing-based approaches (e.g., integron cassette sequencing or metagenomics) to assess physical linkage between *intI1*-associated elements and key ARGs.

## Data Availability

Protocols and data used in this paper are available at a dedicated data repository at Open Science Framework (OSF.io). DOI: 10.17605/OSF.IO/BTZHE

https://osf.io/btzhe/.

## Notes

### Competing Interest Statement

The authors have declared no competing interest.

### Clinical Trial

NCT02362932

### Funding Statement

This study was funded by the Bill & Melinda Gates Foundation (www.gatesfoundation.org) Grant (OPP1137224 to JB) and by USDA NIFA (2025-68015-44818 to DC). The funders had no role in study design, data collection and analysis, decision to publish, or preparation of the manuscript.

